# Chronotype changes after sex hormone use: a prospective cohort study in transgender users of gender-affirming hormones

**DOI:** 10.1101/2023.06.15.23291434

**Authors:** Margot W. L. Morssinkhof, Annefleur Zwager, Karin van der Tuuk, Martin den Heijer, Ysbrand D. van der Werf, Dirk Jan Stenvers, Birit F. P. Broekman

## Abstract

**Background:** Chronotype, an individual’s preferred sleep-wake timing, is influenced by sex and age. Men report a later chronotype than women and age is associated with earlier chronotype. The sex-related changes in chronotype coincide with puberty and menopause. However, the effects of sex hormones on human chronotype remain unclear.

**Aim:** To examine the impact of 3 months of gender-affirming hormone therapy (GAHT) on chronotype in transgender persons.

**Methods:** This study used data from 93 participants from the prospective RESTED cohort, including 49 transmasculine (TM) participants starting testosterone and 44 transfeminine (TF) participants starting estrogens and antiandrogens. Midpoint of sleep and sleep duration were measured using the ultra-short Munich ChronoType Questionnaire (µMCTQ).

**Results:** After 3 months of GAHT, TM participants’ midpoint of sleep increased by 24 minutes (95%CI: 3 to 45), whereas TF participants’ midpoint of sleep decreased by 21 minutes (95%CI: −38 to −4). Total sleep duration did not change significantly in either group.

**Conclusion:** This study provides the first prospective assessment of sex hormone use and chronotype in transgender persons, showing that GAHT can change chronotype in line with cisgender sex differences. These findings provide a basis for future studies on biological mechanisms and clinical consequences of chronotype changes.

## Introduction

The human circadian timing system regulates 24-hour rhythms in behaviour and physiological processes. These circadian rhythms are orchestrated by the central brain clock located in the suprachiasmatic nucleus (SCN) of the hypothalamus, which synchronizes a multitude of peripheral clocks throughout the body. The molecular mechanism of the central and peripheral clocks is the transcriptional translational feedback loop of the core clock genes, which has an intrinsic period duration of approximately 24 hours. The molecular clocks regulate the timing of output genes and thus the timing of physiological processes in specific tissues. In the absence of external time cues, the endogenous circadian cycle lasts approximately 24 hours (Roenneberg, Daan, et al., 2003). The duration of the diurnal rhythm of the central clock can, however, differ between individuals due to various factors (e.g. genetics, environmental factors, biological differences), causing some individuals to have circadian rhythms significantly longer or shorter than 24 hours, resulting in a propensity to sleep earlier or later (Roenneberg et al., 2019).

This preference in sleep-wake timing based on the intrinsic circadian rhythm is also known as “chronotype” (Adan et al., 2012). People with early chronotypes, ("morning types") rise early, have earlier diurnal peaks in physical and mental performance, and retire early in the evening. Conversely, people with a later chronotype ("evening types") are more inclined to have their time peak activity in the second half of the day and preferably go to bed later (Adan et al., 2012).

Previous research has established that people with a late chronotype have a higher risk of sleep problems and poorer mental and physical health (K. S. Jankowski et al., 2019). One potential explanation is that evening-types are more likely to experience sleep loss because they prefer later bed-and wake-up times, but must wake up early to fulfill social obligations. This results in a longer so-called “social jetlag”. Social jetlag is a form of circadian misalignment, caused by a discrepancy between biological sleep timing preferences and social rhythms (K. Jankowski, 2015; Wittmann et al., 2006). Recognizing social jetlag is important since it is associated with an adverse endocrine, behavioral and cardiovascular risk profile due to circadian misalignment and a chronic lack of sleep (Baron & Reid, 2014; Caliandro et al., 2021; Rutters et al., 2014).

The variability in human chronotype is mostly determined by genetic variation in clock genes (Archer et al., 2003; Toh et al., 2001; Vink et al., 2001) and environmental factors (e.g. light exposure) (Montaruli et al., 2021; Roenneberg, Wirz-Justice, et al., 2003). Additionally, research has also showed differences in chronotype based on age and sex. For example, children exhibit a morning preference and develop a propensity towards later chronotype during puberty. Around the age of 20, there is a peak "lateness", where the chronotype slowly returns back to morning preference with increasing age (Roenneberg et al., 2004).

Sex differences in chronotype emerge at the onset of puberty when increasing levels of sex hormones trigger the development of secondary sex characteristics. It is known that girls tend to go into puberty earlier than boys, and the shift to the puberty-associated late chronotype also happens one year earlier in girls than it does in boys (Hagenauer & Lee, 2012). During puberty, the shift to a later chronotype is more pronounced in boys than in girls (Fischer et al., 2017). This difference remains during the reproductive age: adult men generally tend to have a later chronotype than adult women (Adan & Natale, 2002). The disparity in chronotype between men and women disappears around 40 years of age, coinciding with the perimenopause in women (Fischer et al., 2017; Randler & Engelke, 2019). A similar pattern is seen in reported sleep duration: after adolescence, women report a longer sleep duration than men (Kocevska et al., 2021), and these sex differences in sleep duration disappear during the perimenopause (Tonetti et al., 2008).

The changes in chronotype during puberty and menopause have led researchers to hypothesize that sex hormones could be contributing to shifts in chronotype. This could be explained by the presence of estrogen receptors and androgen receptors in the SCN, which are expressed in a sex-specific way (Kruijver & Swaab, 2002). These also have effects on the circadian system: both androgens and estrogens could affect photic sensitivity within the entrainment pathway in rodents, meaning they can moderate the effect of light-and dark cues on the SCN and thereby modify the circadian system in rodents (Joye & Evans, 2022).

Thus far, human research has mainly focused on the association between sex hormones and insomnia (Morssinkhof et al., 2023), while studies on sex hormones and chronotype are scarce. Studies that have examined the association between sex hormones and chronotype show indications that higher testosterone is associated with later chronotypes. Randler et al. (2012) and Jankowski et al. (2019) both found associations between testosterone levels and later chronotypes in males. However, Yuan *et al*. (2023) found that only in women, and not in men, free testosterone was associated with a later chronotype. These studies all assessed cross-sectional associations, and it is therefore still unknown whether the sex hormones could also have a causal effect on the sex differences in chronotype in humans.

Transgender individuals who use gender-affirming hormone therapy (GAHT) are a unique group in whom we could prospectively study effects of sex hormone use on chronotype. Transmasculine (TM) persons, who were assigned female at birth and desire masculinization, can use testosterone, whereas transfeminine (TF) persons, who were assigned male at birth and desire feminization, can use estrogens and anti-androgens. GAHT has as an effect on multiple homeostatic bodily processes and causes significant changes in physical appearances towards the phenotype of the other sex (Cocchetti *et al.,* 2022), but their effects on chronotype are not yet known.

This study aims to examine the effect of sex hormones on chronotype, specifically the midpoint of sleep and sleep duration, in transgender persons after 3 months of GAHT use. We hypothesize that TM persons, using testosterone, show a change in chronotype from early to later-type and a shorter sleep duration after 3 months of GAHT, whereas TF persons, using estrogen and anti-androgens, show a chronotype moving from a later to earlier chronotype and a longer sleep duration after 3 months of GAHT.

## Methods

### Study population

Participants for this study were recruited from the Relationship between Emotions and Sleep in Transgender persons: Endocrinology and Depression (RESTED) study, a prospective cohort study investigating the effects of gender-affirming sex hormone use on sleep and mood. RESTED participants were recruited at the gender clinics of the Amsterdam University Medical Centers (Amsterdam UMC) and the University Medical Centre Groningen (UMCG). In the RESTED study, adults were eligible for participation if they were aged between 18 and 50, diagnosed with gender dysphoria and/or gender incongruence, and planning to start the use of gender-affirming hormone therapy (GAHT).

Exclusion criteria for the RESTED study included: having a pre-existent sleep disorder, use of sleep medication (benzodiazepines, barbiturates, and opiates), or previous use of gender-affirming hormones. Individuals interested in participating in the study received oral and written information about the study protocol, after which informed consent was obtained. The RESTED study was classified as a ‘non-WMO’ study by the Medical Ethical Committee of the Amsterdam UMC (location VUmc) and the local committee at the UMCG, meaning that the Medical Research Involving Human Subjects Act (WHO) did not apply to the data collection of this study (study id. 2019.353).

### Data collection

The study data was collected at the Amsterdam UMC and UMCG between December 2019 and January 2023. A total of 99 participants was included in the RESTED study, of whom 51 TM participants, who were assigned female at birth and started masculinizing GAHT, and 48 TF participants, who were assigned male at birth and started feminizing GAHT. Participants provided measurements before starting GAHT, after 3 months of GAHT, and after 12 months of GAHT. The study measures at every timepoint included questionnaires on depressive symptoms, using the IDS-SR (Rush et al., 1996), stress, using the Perceived Stress Scale (Cohen et al., 1983), sleep quality, using the Pittsburgh Sleep Quality index (Buysse et al., 1989), and insomnia, using the Insomnia Severity Index (Bastien et al., 2001), as well as seven nights of ambulatory sleep EEG measurements, using an ambulatory single-electrode EEG sleep measurement device (Smartsleep, Philips, the Netherlands). The results of these aforementioned assessments are not reported on here. Chronotype was studied using the ultra-short Munich ChronoType Questionnaire (µMCTQ) (Ghotbi et al., 2020).

For the current study, data from 97 participants with available complete chronotype questionnaires at baseline and 3 months were included. Participants were excluded if their chronotype questionnaires were missing at both measurements or, in the case of TF participants, if they used only anti-androgens or only estrogens instead of both. This resulted in the sample sizes as displayed in Figure 1.

**Figure 1.**
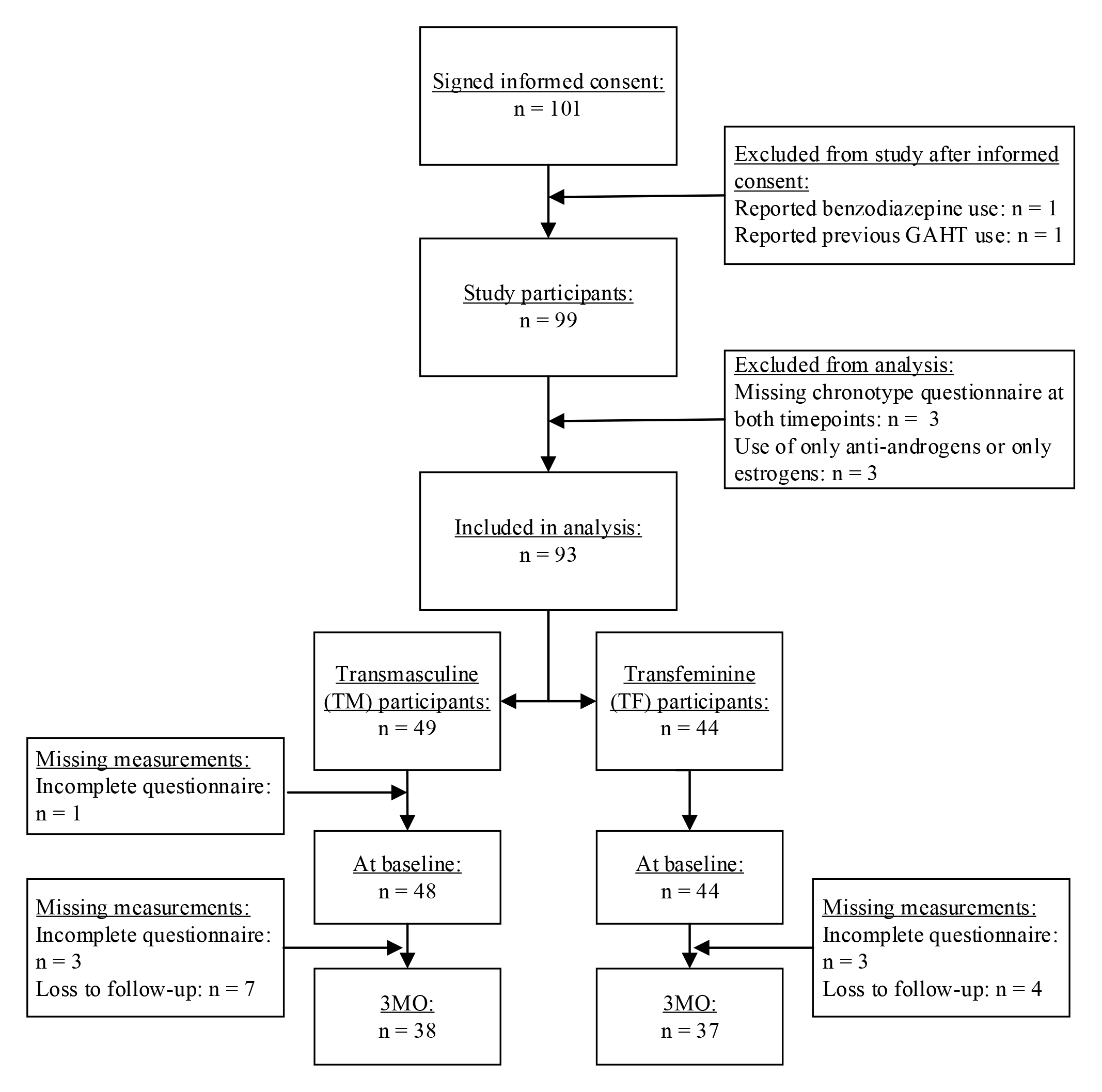
Flowchart in-and exclusions in the RESTED study, sample sizes per group and per measurement timepoint and drop-outs and missing measurements per timepoint and group.

### Treatment protocol

For the TM participants, gender-affirming hormone therapy consisted of testosterone and, for some, additional use of cycle-regulating medication (e.g. progestins or hormonal contraceptives). Testosterone was administered as either transdermal gel (50 mg once daily), intra-muscular injection of short-acting testosterone esters (250 mg once per 3 weeks) or as an intra-muscular injection of long-acting testosterone undecanoate (1000 mg once every 12 weeks). Participants could also use medication to regulate their menstrual cycle, either using progestogens (lynestrenol, norethisteron, levonorgestrel, medroxyprogesterone) or combined estrogen-progestin oral contraceptives (COCs).

For the TF participants, gender-affirming hormone therapy consisted of estrogens and anti-androgens. Estradiol was either administered orally, using estradiol valerate (2 mg twice daily), or transdermally with an estradiol patch (100 µg patches every 3 days) or estradiol gel (1 – 1,5 mg once daily). Anti-androgens were used in the form of cyproterone acetate (10 mg daily) or injections of the GnRH analog triptorelin (3.75 mg once every 4 weeks or 11.25 mg every 12 weeks) or leuproreline (3.75 mg once every 4 weeks). Several TF participants started using only anti-androgens (n = 1) or estrogens (n = 2) at the start of GAHT: these participants were excluded from the current analyses. A flowchart of participant in-and exclusions is displayed in figure 1.

### Demographic and clinical characteristics

Participants visited the clinic at the start and after 3 months of GAHT, and clinical characteristics (e.g. sex assigned at birth, age at baseline measurement, body mass index, medication use, form and dosage of hormone use, use of menstrual cycle regulation or contraceptives) were obtained from the medical files from these appointments.

At the Amsterdam UMC and UMCG, serum testosterone measurements were conducted using liquid chromatography-tandem mass spectrometry (LC-MS/MS) with a lower limit of quantitation of 0.1 nmol/L, and an inter-assay coefficient of variation of 4% to 9%. Serum estradiol measurements were conducted using LC-MS/MS with a lower limit of quantitation of 20 pmol/L and an inter-assay coefficient of variation of <7%.

### Chronotype

The primary outcome of this study was change in chronotype, measured by changes in midpoint of sleep at baseline and after 3 months of GAHT and by changes in sleep duration at baseline and after 3 months of GAHT.

The sleep-corrected Midpoint of Sleep on Free days (MSF_sc_) and sleep duration on free days were measured using the ultra-short Munich ChronoType Questionnaire (µMCTQ) (Ghotbi et al., 2020). The µMCTQ is a validated short questionnaire used to determine sleep-wake-behavior in a regular week, containing questions about sleep onset and wake times on workdays and work-free days, use of an alarm clock on free days, number of workdays, and doing shift work. We used two main outcomes from the µMCTQ: reported sleep duration and MSF_sc_. Additionally, we used the items on the number of work days one has in a week and use of an alarm clock on free days.

Chronotype is determined based on sleep timing on work-free days, since these are assumed to be relatively free of constraints on sleep-wake behavior, such as an early work schedule. Chronotype is typically represented as the mid-point of sleep on work-free days (MSF), which is calculated based on the midpoint between the time of sleep onset and the time of sleep end on work-free days. Since people with late chronotypes tend to accumulate sleep debt throughout the week, the chronotype can be corrected for the accumulated sleep debt, creating the “sleep debt corrected” midpoint of sleep on work-free days (or MSF_sc_). The MSF_sc_ is calculated by weighing the average sleep duration on work days compared to the sleep duration on work-free days for people who sleep longer on work-free days than on work days. In people who do not sleep longer on work-free days than on work days, the MSF equals the MSF_sc_. Sleep duration is the weighted average of the time between sleep onset and sleep offset on work days and on work-free days, corrected by the ratio of work to work-free days.

### Statistical analyses

R studio (version 4.0.3) was used for all statistical analyses. Data analysis was performed using linear mixed effect models using the R packages lme4 (Bates et al., 2015) and lmerTest (Kuznetsova et al., 2017). To account for repeated measures in the same person, a random intercept was used for each subject. Analyses were conducted separately in the TM and TF groups.

To estimate the changes in midpoint of sleep and sleep duration after 3 months of GAHT, we used the measurement time point (e.g. baseline or 3-month follow-up) as a fixed predictor in the unadjusted model, as displayed below. Secondly, we incorporated a possible confounder (work status: working more or less than 3 days a week) into account in the adjusted model. Thirdly, to account for the use of alarm clocks in the cohort, we conducted a sensitivity analysis where we included only the subgroup of the cohort who reported not using an alarm clock on free days. All models are displayed below.

**Unadjusted model**: Outcome ^a.^ ∼ measurement time point + (1|Participant ID)

**Adjusted model**: Outcome ^a.^ ∼ measurement time point + Work status ^b.^ + (1|Participant ID)

**Sensitivity model in subgroup _c._**: Outcome ^a.^ ∼ measurement time point + Work status^b.^ + (1|Participant ID)

a. The following outcomes were tested: sleep duration, MSF_sc_
b. Work status: whether participant reports working or going to school 3 or more days per week or working or going to school less than 3 days per week.
c. This model was conducted on a subgroup of participants who reported not using an alarm on free days.

## Results

### Demographic characteristics

The sociodemographic characteristics of the entire study sample at baseline and after 3 months of GAHT are reported in Table 1. At baseline, TM participants had a median age of 23 ± 5 years and 63% had more than 3 days of work or school per week. At the start of GAHT, testosterone gel was the most commonly utilized type of GAHT in TM participants (90%), short-acting testosterone injections were used by the rest of the group (10%), and 44% of TM participants used cycle regulation medication, of whom 23% a form containing only progestogens and 21% a combined oral contraceptive with estradiol and progestogens.

**Table 1.** Characteristics of the transmasculine (TM) and transfeminine (TF) groups reported per measurement time point.

|  | *TM participants* |  | *TF participants* |  |
| --- | --- | --- | --- | --- |
| *Measurement point* | *Baseline*<br>(*n* = 48) | *3 months*<br>(*n* = 38) | *Baseline*<br>(*n* = 44) | *3 months*<br>(*n* = 37) |
| *Age (years; median, interquartile range)* | 22 (19.8; 24) | 22 (19.5; 24) | 26 (24; 31) | 27 (24.8; 32.3) |
| *Alcohol use* |  |  |  |  |
| *Units per week (median, interquartile range )* | 0 (0; 1) | 0.5 (0; 0.5) | 0.5 (0 to 2) | 0.5 (0 to 2) |
| *Missing* | 1 | 13 | 2 | 12 |
| *Smoking (n, %)* |  |  |  |  |
| *Current smoker* | 5 (10%) | 3 (8%) | 6 (14%) | 1 (3%) |
| *Not current smoker* | 42 (88%) | 25 (66%) | 38 (67%) | 31 (84%) |
| *Missing* | 1 (2%) | 10 (26%) | 0 (0%) | 5 (13%) |
| *Use of alarm clock on free days (n, %)* | 17 (35%) | 14 (35%) | 15 (34%) | 12 (32%) |
| *Work or school days per week (n, %)* |  |  |  |  |
| *Less than 3 days of school or work* | 18 (37%) | 11 (29%) | 19 (43%) | 19 (51%) |
| *3 or more days of school or work* | 30 (63%) | 27 (71%) | 25 (57%) | 18 (48%) |
| *Serum hormone levels (median, interquartile range) <sup>a</sup>.* |  |  |  |  |
| *Testosterone (nmol/L)* | 0.9 (0.6; 1.2) | 19 (13.5; 24.5) | 13 (10; 17.5) | 0.55 (0.4; 0.8) |
| *Estradiol (pmol/L)* | 114 (79; 208) | 134 (104; 186) | 71.5 (59.8; 89.5) | 283 (180; 396.3) |

<sup>a</sup>. Reference values for cisgender men: testosterone, 9 to 30 nmol/L, estradiol, 12 pmol/L to 126 pmol/L. Reference values for premenopausal cisgender women: testosterone, 0.3 to 1.6 nmol/L, estradiol, 31 pmol/L to 2864 pmol/L. (Amsterdam UMC Endocrinologisch Lab et al., z.d.; Verdonk et al., 2019)

At baseline, TF participants had a median age of 27 ± 5 years and 43% had school or work on more than 3 days per week. The most common form of estradiol prescribed at the start of GAHT was oral estradiol (61%), the second most common form were estradiol patches (30%) and 9% used estradiol gel. The majority of TF participants started using a form of GnRH-analogues as testosterone suppressant (84%) at the start of GAHT, and three TF participants started using cyproterone acetate (9%).

### Transmasculine group

As displayed in table 2 and in figure 2, the TM group shows a 20-minute later MSF_sc_ after 3 months of masculinizing GAHT in the unadjusted model, a 22-minute later MSF_sc_ in the adjusted model and a 30-minute later MSF_sc_ in the sensitivity analysis. The TM group shows no changes in the sleep duration after 3 months of GAHT in the unadjusted or adjusted model. In the sensitivity analysis, the TM group shows a trend towards a longer sleep duration: the sleep duration is estimated to be 27 minutes longer (p = 0.054) in TM users who did not use an alarm clock on free days. All results from the TM group are displayed in table 2 and figure 2.

**Figure 2.**
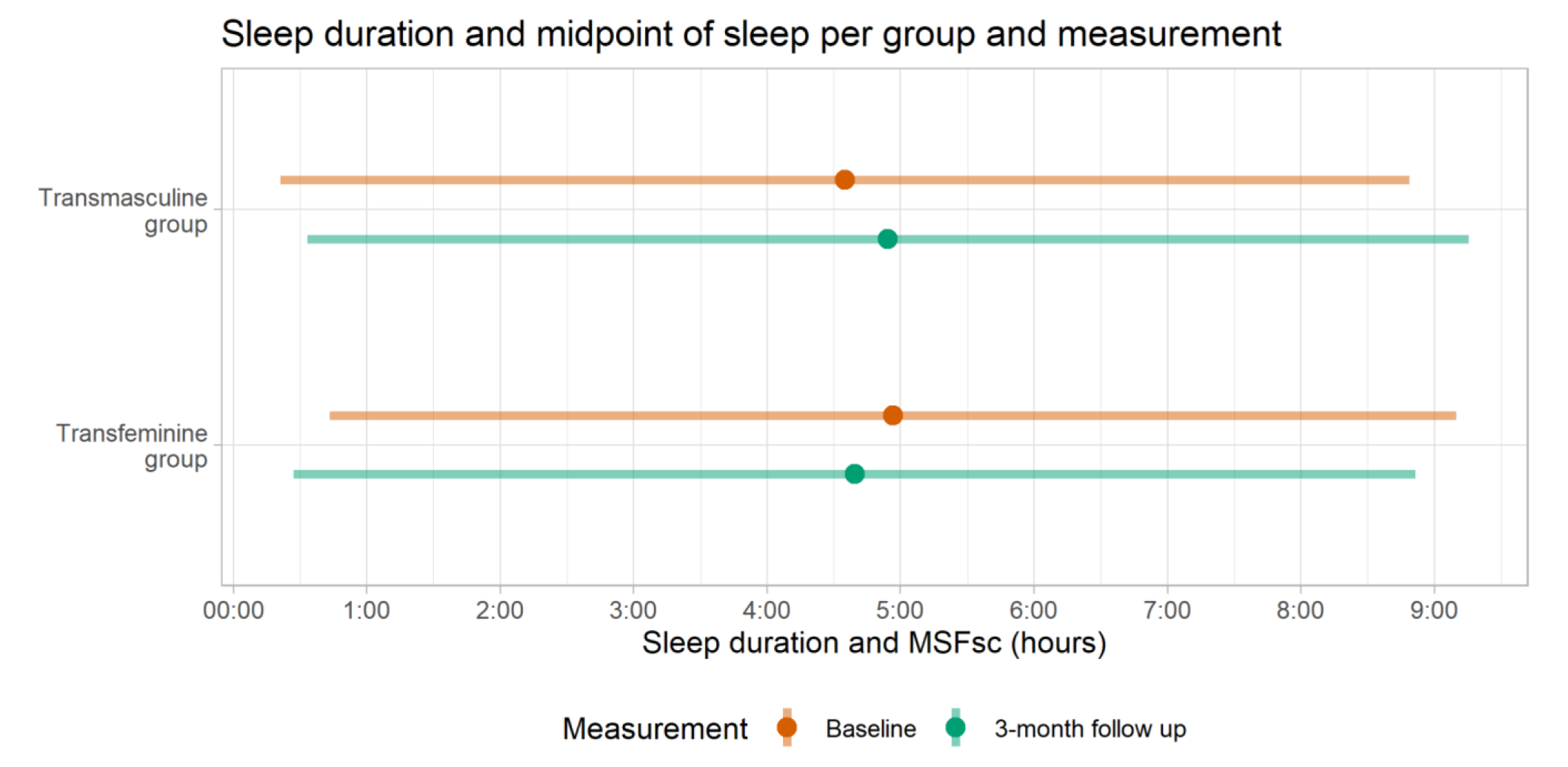
Sleep duration (line) and sleep-corrected midpoint of sleep on free days (point) before GAHT (baseline; displayed as the top orange bar) compared to after 3 months of GAHT (displayed as the bottom green bar). The Y-axis displays the groups, the X-axis displays the sleep duration (as a line) and midpoint of sleep (as a point) in hours.

**Table 2.** Sleep duration and (sleep-corrected) Midpoint of Sleep on Free days (MSF_sc_) obtained from the µMCTQ in the transmasculine (TM) group and the transfeminine (TF) group. All variables are reported in clock hours (hh:mm).

| **Outcome**<br>**in hh:mm** | **Transmasculine group** |  |  |  |  |
| --- | --- | --- | --- | --- | --- |
|  | *Predictor (n)* | *Mean (SD)* | **Unadjusted model**<br>*Estimated change from baseline (95% CI: L; H) p-value)* | **Adjusted model**<br>*Estimated change from baseline (95% CI: L; H) p-value)* | **Sensitivity model <sup>1.</sup>**<br>*Estimated change from baseline (95% CI: L; H) p-value)* |
| **MSF<sub>sc</sub>** | Baseline (48) | 04:35 (01:26) | Reference | Reference | Reference |
|  | 3-month follow up (38) | 04:54 (01:37) | 00:20 (-00:02; 00:41)<br>p = 0.084 | 00:24 (00:03; 00:45)<br>p = 0.03 | 00:30 (00:03; 00:58)<br>p = 0.039 |
| **Sleep duration** | Baseline (48) | 08:28 (01:14) | Reference | Reference | Reference |
|  | 3-month follow up (38) | 08:42 (01:04) | 00:13 (-00:08, 00:35)<br>p = 0.22 | 00:14 (-00:08, 00:36)<br>p = 0.22 | 00:27 (00:01; 00:55)<br>p = 0.054 |
| **Outcome**<br>**in hh:mm** | **Transfeminine group** |  |  |  |  |
|  | *Time point, months after start GAHT (n)* | *Mean (SD) or median (IQR) <sup>1.</sup>* | **Unadjusted model**<br>*Estimated change from baseline (95% CI: L; H) p-value)* | **Adjusted model**<br>*Estimated change from baseline (95% CI: L; H) p-value)* | **Sensitivity model <sup>1.</sup>**<br>*Estimated change from baseline (95% CI: L; H) p-value)* |
| **MSF<sub>sc</sub>** | Baseline (44) | 04:56 (01:27) | Reference | Reference |  |
|  | 3-month follow up (37) | 04:39 (01:02) | -00:15 (-00:35; 00:05)<br>p = 0.14 | -00:21 (-00:38; -00:04)<br>p = 0.023 | -00:17 (-00:32; -00:03)<br>p = 0.03 |
| **Sleep duration** | Baseline (44) | 08:26 (01:05) | Reference | Reference |  |
|  | 3-month follow up (37) | 08:25 (00:54) | 00:00 (-00:21; 00:21)<br>p = 0.99 | -00:02 (-00:22; 00:17)<br>p = 0.83 | -00:02 (-00:22; 00:17)<br>p = 0.86 |

<sup>1.</sup> Excluding participants who use alarm clocks on free days. Sample sizes are $n = 31$ at baseline and $n = 24$ at 3 month-follow up for the TM group and $n = 29$ at baseline and $n = 25$ at 3 month-follow up for the TF group.

### Transfeminine group

In the TF group, the adjusted model for MSF_sc_ showed 21-minute earlier MSF_sc_ after 3 months of GAHT compared to baseline. The sensitivity analysis showed a significant 17-minute earlier MSF_sc_ in TF participants who did not use an alarm clock on free days. The TF group showed no significant changes in sleep duration after 3 months of GAHT. All results from the TF group are also displayed in table 2 and figure 2.

## Discussion

With this study we prospectively investigated the effects of sex hormone use on chronotype in transgender GAHT users. Our results show the transmasculine participants, who were assigned female at birth, developed a later chronotype after the first 3 months of testosterone. In contrast, transfeminine participants, who were assigned male at birth, developed an earlier chronotype after 3 months of estrogens and anti-androgens. Taken together, our results indicate that the use of masculinizing or feminizing sex hormones can influence midpoint of sleep in an opposite direction, which is in line with chronotype differences seen in the cisgender population.

Our findings of a later chronotype after 3 months of masculinizing sex hormone use and an earlier chronotype after 3 months of feminizing sex hormone use are in concordance with our hypotheses. Previous research found that testosterone was associated with a later chronotype: salivary testosterone levels in cisgender men were found to be associated with a later chronotype (Randler et al., 2012), and women with PCOS, who have elevated testosterone levels, also have a later chronotype than women without PCOS (Karasu et al., 2021). Estrogen had the opposite effect: previous research found that estrogen has a phase-advancing effect in rodents, meaning chronotype shifts to an earlier preference (Albers et al., 1981; Leibenluft, 1993). Thus, previous research indicated that the effects of estrogen and testosterone on chronotype might be opposing, which is in line with our current findings. Based on our findings it is not possible to separately assess the role of testosterone and estrogen in the reported chronotype changes. All TF participants used both estrogens and anti-androgens, and therefore the resulting earlier chronotype could both be caused by the increase in estrogen signaling, the decrease in testosterone signaling, or by an interaction of both factors. A similar limitation is found in the TM participants: although they all start using testosterone and the serum testosterone levels strongly increase, many also report cessation of their menstrual cycle, which most likely means that the endogenous estradiol fluctuations have also stopped. Furthermore, some of the TM participants used cycle regulation, which also affects gonadal hormone levels. Therefore, although our results do show that exogenous use of gender-affirming hormones changes chronotype in line with cisgender sex differences, it is not possible to assess which hormonal mechanisms are underlying these shifts in chronotype.

Concerning sleep duration, no changes were found in the TF participants after 3 months of GAHT. This is in contrast with the findings of Liu et al., (2003), who found that men show a sleep duration reduction of approximately 1 hour after administration of a high dosage of testosterone, and Sakaguchi et al., (2006) who discovered that short sleepers had higher testosterone levels. Furthermore, we did not find any change in sleep duration in the TF participants after 3 months of hormone therapy with estrogens and antiandrogens. These findings may indicate that hormone therapy with either testosterone or estrogen and antiandrogens might not directly affect reported sleep duration.

The strengths of this study lie mainly in its unique study population and its prospective study setup. Firstly, the study of transgender hormone users enables us to study the effects of exogenous sex hormones, which are administered in such a dosage that the sex hormone levels in our participants transition from the levels found in cisgender women towards the levels found in cisgender men, or vice versa, as shown in Table 1. This is a unique and novel way of studying effects of sex hormones on chronotype in hormone users. Secondly, the prospective setup enables within-person comparisons of chronotype and sleep duration, meaning the resulting estimates are more reliable for assessing possible causal effects of sex hormones.

A methodological limitation of this study is that a subgroup of participants in this study used alarm clocks on free days. It is preferable to determine someone’s natural rhythm on free days, without the interference of an alarm clock. Therefore, participants who use an alarm clock on free days should be excluded from the calculation of MSF_sc_. The number of participants using an alarm clock on free days was quite large in our cohort, and excluding these subjects would significantly reduce the size of our study sample. Therefore, we decided not to exclude participants who used an alarm clock on free days. We have instead opted to conduct a sensitivity analysis for participants who did not use an alarm clock on free days. We found a 30-minute later MSF_sc_ in the TM group and a significant 17-minute earlier MSF_sc_ in the TF group, indicating that including participants who use alarm clocks on free days did not change the direction of the found effects.

Another limitation in our study setup is that starting GAHT is associated with changes in depression and anxiety, since starting the use of gender-affirming hormones was found to most likely reduce depressive symptoms in both TM and TF groups (Costa & Colizzi, 2016). Depression is related to disturbed sleep and circadian rhythm alterations, meaning that the interaction between depression and chronotype after starting GAHT could be affecting our results (Dai & Hao, 2019; Soria & Urretavizcaya, 2009). Therefore, our results should be interpreted taking into account that participants’ depressive symptoms might change after the start of GAHT. Future research that examines sleep timing in interaction with depressive and anxiety symptoms in transgender persons during GAHT use could contribute to a better understanding of our findings.

The final limitation is that chronotype and related sleep variables were measured through a self-reported questionnaire. Participants estimate their own sleep patterns, which might be an over-or underestimation since only two questions about sleep onset and wake-up time are included in the questionnaire. There is a well-known paradox in sleep and sex differences, showing that cisgender women are more likely to report insomnia and poor sleep, whereas their sleep architecture tends to show longer sleep durations and better sleep compared to cisgender men (Roehrs et al., 2006; Zhang & Wing, 2006). One of the explanations is that sleep perception is different in men and women, and this difference in sleep perception might also be present in the current study. Therefore, in future research more specific questions could be implemented to determine actual sleep time and sleep timing could also be measured objectively using actigraphy measurements.

Overall, our findings show novel evidence for an effect of sex hormones on chronotype. These findings bring up new questions, both on fundamental underlying mechanisms as well as on clinical and behavioral effects of these chronotype changes. Future studies should focus on studying the effects of sex hormones on circadian rhythmicity in the body, through assessment of actigraphy, body temperature, or levels of cortisol and melatonin, to address the underlying mechanisms between sex hormones and chronotype. Furthermore, it should focus on the clinical consequences of changes in chronotype, such as changes in health risk and mental health.

## Data Availability

Data of the present study are available upon reasonable request.

## Acknowledgements

We would like to thank all RESTED study participants, without whom this study would not have been possible. Furthermore, we would like to thank all clinicians of the UMCG and Amsterdam UMC for their collaboration and efforts for the RESTED study. Lastly, we would like to thank all interns and students who assisted in the data collection of the RESTED study: Rens Voskuijlen, Lotte van der Kolk, Claartje Binnerts, Jip Andriesse, Danique Hofs, Laura Varkevisser, Maureen González, Sigrid Theunis, Berend Slootman, Nina Pot and Isabel Löwensteijn.

## Disclosure of interest

This work was supported by the NWO, Netherlands [Veni grant, grant number 91619085, 2018] supplied to BB. The NWO had no involvement in the study design, data collection, analysis or interpretation of the data, or writing of the report. All other authors have no financial or non-financial conflict of interest.

## Data availability statement

Data available on request

## Notes

### Competing Interest Statement

The authors have declared no competing interest.

### Funding Statement

This work was funded by the NWO, Netherlands [Veni grant, grant number 91619085, 2018] supplied to BB. The NWO had no involvement in the study design, data collection, analysis or interpretation of the data, or writing of the report.

### Author Declarations

The RESTED study was classified as a non-WMO study by the Medical Ethical Committee of the Amsterdam University Medical Centers (location VUmc) and the local committee at the University Medical Center Groningen, meaning that the Medical Research Involving Human Subjects Act (WHO) did not apply to the data collection of this study (study id. 2019.353).

